# A Genome-Wide Association Study Reveals Novel Genetic Markers Associated with Human African Trypanosomiasis

**DOI:** 10.1101/2024.05.14.24307227

**Authors:** Julius Mulindwa, Magambo Phillip Kimuda, Harry Noyes, Hamidou Ilboudo, Mathurin Koffi, Bernadin Ahouty, Oscar Nyangiri, Anneli Cooper, Caroline Clucas, Peter Nambala, Janelisa Musaya, Dieudonne ́ Mumba Ngoyi, Kevin Karume, Olivier Fataki, Gustave Simo, Elvis Ofon, John Enyaru, Barbara Nerima, Andy Tait, Lucio Marcello, John Chisi, Jacques Kabore, Justin Windingoudi Kabore, Kelita Kamoto, Martin Simuunza, Vincent P. Alibu, Vincent Jamonneau, Marianne Camera, Mamadou Camara, Bruno Bucheton, Christiane Hertz-Fowler, Annette Macleod, Enock Matovu, TrypanoGEN Research Group as members of The H3Africa Consortium

**Author notes:** To whom correspondence should be sent. Currently at Institut de Recherche en Sciences de la Santé, Unité de Recherche Clinique de Nanoro (IRSS/URCN), Burkina Faso. Currently at Biomathematics and Statistics Scotland, Edinburgh (Lucio Marcello). Currently at Wellcome Trust, London, UK. These authors contributed equally to the work (Julius Mulindwa, Magambo Phillip Kimuda, Harry Noyes).

## Abstract

*Trypanosoma brucei gambiense* and *Trypanosoma brucei rhodesiense* cause human African trypanosomiasis (HAT), a neglected tropical disease that constitutes an important public health issue in sub-Saharan Africa. In the absence of a vaccine, only chemotherapy and vector control has been used to combat the disease. Environmental factors, such as exposure to infected tsetse files, and genetic factors such as variants in the *APOL1* gene have been shown to contribute to the risk of developing HAT. However, the known factors only explain a small part of the risk of developing trypanosomiasis. We have undertaken a genome wide association study (GWAS) using 3813 samples from *T. b. gambiense* and *T.b. rhodesiense* HAT foci in Guinea, Côte d’Ivoire, Cameroon, DRC, Malawi and Uganda. 2141 samples were genotyped on the H3Africa SNP chip followed by a genotyping a validation cohort of an additional 1,627 samples at candidate loci. After the primary and validation studies we identified a novel locus near *SMOC2* with genome-wide significance. We also identified suggestive associations near *NXN, NTNG1* and *NCKAP5* that have stronger associations with disease susceptibility than the *APOL1* loci that has been previously identified by hypothesis driven approaches. These genes offer new entry points for future studies of the underlying genetic mechanisms of HAT.

## Introduction

Human African Trypanosomiasis (HAT) is caused by two subspecies of *Trypanosoma brucei*, namely *T. brucei rhodesiense* that causes the acute form of the disease while *T. b. gambiense* causes the chronic form of the disease. HAT is transmitted by the bite of an infected tsetse fly. The disease in mammalian hosts manifests in two stages, the hemolymphatic and meningoencephalitic stages. While over the last decade there has been a steady decline in the number of sleeping sickness cases reported in sub-Saharan Africa (Franco et al. 2024; 2022)(Dickie et al. 2020) HAT continues to be a public health concern in Sub-Saharan Africa. Despite the efforts to control the disease there are still sporadic epidemics such as the unexpected outbreaks of Acute HAT in Malawi in 2019-2020 (Nambala et al. 2022) and Ethiopia in 2022 (Abera et al. 2024). Reported cases only represent a fraction of the true prevalence (Büscher et al. 2018) and the growing realization that HAT has a human reservoir of asymptomatic carriers indicates that there is an ever-present risk of new HAT epidemics, even in the presence of surveillance and control programmes.

Whilst HAT has traditionally been considered to be invariably fatal, asymptomatic individuals represent a group of people that remain infected without symptoms for many years. They are defined as being positive by serology on the card agglutination test for trypanosomiasis (CATT) for several years and having a positive trypanolysis test but are negative by parasitology in blood or lymph (Jamonneau et al 2012, Büscher et al. 2018). Recent studies have shown that many of these individuals carry parasites in their skin and form an important reservoir of infection that could play a role in sustaining endemicity, particularly for chronic HAT (Capewell et al. 2016; Camara et al. 2021). Analysis of the parasites’ genomes shows very little variation indicating that host genomics could be playing a bigger role in determining susceptibility and/or resistance to infection than parasite variation (Weir et al. 2016).

The TrypanoGEN consortium and others have conducted at least 11 association studies on cytokine and immune related genes (Cooper et al. 2017) (Ofon et al. 2017; 2019; Kimuda et al. 2018; Kaboré et al. 2017; 2019; Fataki Asina et al. 2020; Ahouty et al. 2017)(Kamoto et al. 2019) (Hardwick et al. 2014; Courtin et al. 2007; 2006). These studies have found some interesting associations between genes and risk of HAT in individual populations, notably with *APOL1*. APOL1 is the trypanolytic factor in human serum that kills most trypanosome species. *T.b. gambiense* and *T.b. rhodesiense* both developed independent mechanisms of resistance to APOL1. We have shown that variations in *APOL1* are associated with resistance to *T.b. gambiense* and *T.b. rhodesiense* infections and disease severity. However, these variants are also associated with risk of chronic kidney disease (Genovese et al. 2010; Cooper et al. 2017), and drugs to reduce APOL1 levels are being developed (Egbuna Ogo et al. 2023). Although candidate gene studies have been successful in identifying important functional variants regulating Mendelian conditions, the novel associations that are identified in GWAS are frequently with genes that have not previously been considered likely candidates, leading to new insights into disease mechanisms and new opportunities for intervention. We have therefore undertaken a GWAS to identify novel candidate loci that might regulate response to infection with African trypanosomes. This can eventually lead to the discovery of the pathways that are most important in regulating response to infection and the identification of novel therapeutic opportunities including repurposing existing drugs for use in HAT.

## Materials and Methods

### Ethical compliance (consent)

Before individuals could participate in this study they were required to give informed written consent. This was provided in English, French, and, where possible, their local languages. In instances where they were unable to read or write a trained clinician and/or ethicist was on hand to help with administering the consent forms. Ethics approval for this study was provided by each of the TrypanoGEN consortium member country ethics review committees (Mulindwa et al. 2020) (Ilboudo et al. 2017). These were as follows: Democratic Republic of Congo (Minister de la Sante Publique: No 1/2013); Côte d’Ivoire (Ministere de la Sante et de la Lutte Contre le SIDA; Uganda (Vector Control Division Research Ethics Committee (Ministry of Health), Uganda National Council for Science and Technology HS 1344); Guinea (Comite Consultatif de Deontologie et d’Ethique [CCDE] de l’Institut de Recherche pour le Developpement: 1-22/04/2013); Cameroon (Le Comité National d’Ethique de la Recherche pour la Santé Humain: 2013/364/L/CNERSH/ SP), Comité National D’Ethique et de la Recherche 2014/No 38/ MSLS/CNER-dkn); Malawi National Health Sciences Research Committee, protocol numbers NHSRC15/4/1399 and Malawi 1213. During the administering of the consent form it was explained to the participants how the biological specimens would be collected, used, and stored. Description of data sharing/access and protection of participant anonymity are described in detail in (Ilboudo et al. 2017).

#### Recruitment

In this GWAS, the study cohort consisted of individuals from both acute (*T.b.rhodesiense*) and chronic (*T.b.gambiense*) HAT endemic regions of sub-Saharan Africa. The cohort included HAT cases and controls as well as individuals described as asymptomatic or latent. Recruitment strategies varied between countries and are described in our smaller scale candidate gene studies (Cooper et al. 2017) (Ofon et al. 2017; 2019; Kimuda et al. 2018; Kaboré et al. 2017; 2019; Fataki Asina et al. 2020; Ahouty et al. 2017)(Kamoto et al. 2019). In Guinea and Côte d’Ivoire samples were actively collected during regular surveillance by teams from the Ministries of Health in the known foci. In DRC and Cameroon samples were mainly passively collected from patients attending clinics for treatment whilst in Uganda and Malawi a mixture of active and passive sampling strategies was used. Case definitions were as previously described (Ilboudo et al. 2017), briefly: cases of *T. b. gambiense* HAT were positive on the card agglutination test for trypanosomiasis (CATT) with a titre ≥ 1/8 and had parasites detectable by microscopy. Mini-anion exchange columns were used for concentration of parasites on samples that were positive by serology. Participants with latent infections were identified in Guinea and Côte d’Ivoire and were serology positive by CATT and the trypanolysis test but no parasites were observed by microscopy. These individuals were followed up for at least two years and remained CATT and trypanolysis test positive. Control samples were collected from participants in the same communities and were negative on both the CATT and trypanolysis test and had no known history of HAT. There is no serological test for *T. b. rhodesiense* infections and cases were identified by microscopy and controls were matched from the same communities and had no signs or symptoms of HAT and were PCR negative for *T. b. rhodesiense* (Kimuda et al. 2018; Kamoto et al. 2019).

### Genotyping and data extraction

DNA was genotyped by Illumina Corp. San Diego on the Infinium H3Africa Consortium Array V1 which contains 2,267,376 SNP (The H3Africa Consortium et al. 2014). Genotypes were extracted from Illumina gtReport files into Plink format files with the topbottom.nf Nextflow workflow (Hazelhust 2018). Genotypes were imputed for 81,694,344 SNP by the H3Africa Imputation service at H3Bionet service using the 1,000 Genomes reference panel (Abecasis et al. 2012) (Sengupta et al. 2023).

88 SNP at 57 loci identified in the primary analysis were genotyped in an additional 1,627 participants samples by LGC Biosearch Technologies, Hoddesden, UK using the KASP™ genotyping assay.

The APOL1 G2 indel is not represented on the H3Africa SNP chip and was genotyped independently in all samples by the same technology.

### Data Quality Control and Filtering strategy for SNPs and study participants

Quality control was carried out using Plink and R. The criteria were as follows: loci that had > 10% missing data or minor allele frequencies (MAF) < 5% or deviations from Hardy Weinberg Equilibrium (p < 0.0001) or heterozygosity > 0.30 or heterozygosity < 0.245 were filtered out. Additionally, loci that were that showed differences in missingness between cases and controls were also filtered.

### GWAS analysis using logistic regression models and linear mixed models

The GWAS was performed using a Linear mixed model (LMM) with a kinship relatedness matrix compiled from a pruned set of unlinked SNP across the whole genome to control for population structure, this was implemented using GEMMA (version 0.98.1) (Zhou and Stephens 2012). An initial whole genome analysis on the genotyped SNP was followed by an analysis of imputed SNP within 100kb of any SNP with a raw p < 10^−5^ in the primary analysis. Imputed SNPs were subject to the same filters as primary SNP and the same relationship matrix was used for the primary analysis on genotyped SNP and the secondary analysis of imputed SNP. Results were visualised using the ggbio package in R (Yin, Cook, and Lawrence 2012).

### Replication

SNP that were genotyped in the validation cohort were tested for association with HAT using the Fisher Exact test in Plink (Purcell et al. 2007). Summary statistics for associations in the primary GWAS and secondary validation cohorts were combined using METAL to obtained final association p values (Willer, Li, and Abecasis 2010).

## Results

### Study populations

A total of 3,813 adult participants between the ages of 3 months and > 75 years old were recruited into the study (Table 1). Participants were classified as cases, controls or latent infections. Cases were defined as participants who had trypanosomes detected in blood, by microscopy. Asymptomatic or latent infections were defined as participants who had positive CATT and trypanolysis tests but had no parasites in blood by microscopy at initial visit and at follow up over at least two years. Latent cases were only recruited in Guinea and Côte d’Ivoire where long-term control programme repeatedly survey the same endemic communities. Controls were defined as individuals who lived in the same areas as cases and were CATT and trypanolysis negative and showed no clinical signs or symptoms indicating current or previous HAT infection. They were also matched for age and sex with HAT cases. After an initial quality control, the samples were divided into a discovery cohort of 2,141 samples, followed by a validation cohort of 1,627. The discovery cohort was genotyped on the H3Africa Illumina chip and the validation cohort was genotyped for selected candidate loci that emerged from the GWAS (Table 1).

**Table 1.** Counts of samples in the discovery and validation cohorts from each population used in the genetic analyses after filtering out low quality samples. Populations were defined by country except in the case of Uganda where there are two populations: one of speakers of Nilotic languages from a *T. b. gambiense* endemic area of northwest Uganda and one of speakers of Niger Congo languages from *T. b. rhodesiense* endemic areas of Central and South East Uganda.

| Population | Phenotype | Discovery Cohort | Validation Cohort |
| --- | --- | --- | --- |
| Cameroon | Case | 8 | 47 |
| Cameroon | Control | 29 | 109 |
| DRC | Case | 143 | 180 |
| DRC | Control | 99 | 482 |
| Guinea | Case | 351 | 100 |
| Guinea | Control | 144 | 112 |
| Guinea | Latent | 66 | 59 |
| Côte d'Ivoire | Case | 66 | 42 |
| Côte d'Ivoire | Control | 101 | 168 |
| Côte d'Ivoire | Latent | 28 | 10 |
| Malawi | Case | 111 | 39 |
| Malawi | Control | 211 | 40 |
| Uganda Tbg | Case | 121 | 86 |
| Uganda Tbg | Control | 188 | 79 |
| Uganda Tbr | Case | 158 | 76 |
| Uganda Tbr | Control | 317 | 43 |

Principal component analysis of GWAS genotype data showed that the genetic relationships between populations are broadly similar to geographic relationships (Fig 1). There is large genetic diversity within Uganda compared to that observed within West Africa (Guinea and Côte d’Ivoire) or Central Africa (Cameroon, DRC and Malawi).

**Figure 1.**
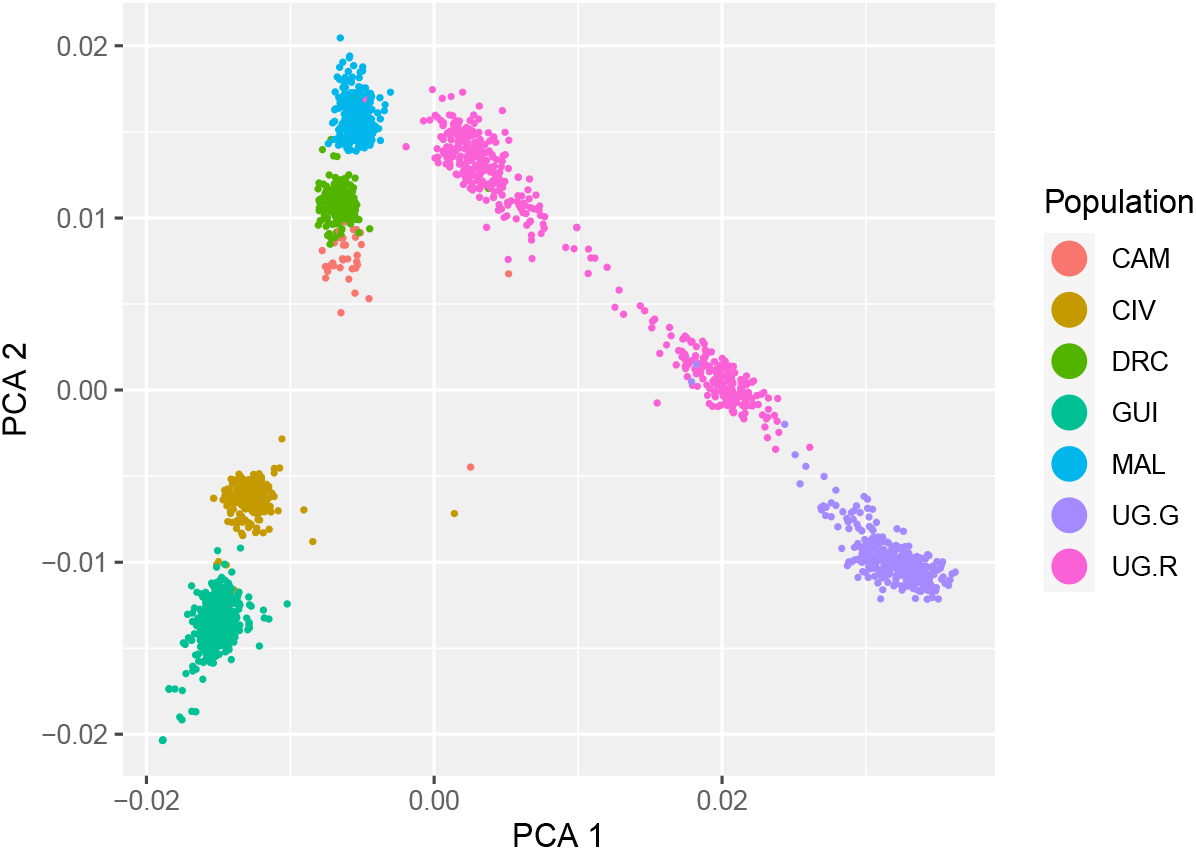
PCA plot showing relationships between participants; participants are coloured by population (CAM = Cameroon, CIV = Cote D’Ivoire, DRC = Democratic Republic of the Congo, GUI = Guinea, Mal = Malawi, UG.G = North West Uganda *T.b. gambiense* endemic area, UG.R = South East and Central Uganda *T.b. rhodesiense* endemic areas) genetic relationships broadly correspond to geographic relationships.

### Two stage study design and power

#### GWAS primary Analysis

Given the three phenotypes available (case, control and latent) and the genetic diversity of both the human and parasite populations, the data were analysed using five contrasts which are shown in Table 2 together with counts of participants in each.

**Table 2.** List of populations used in GWAS analyses, and the counts of participants used in each. Tbg case latent and Tbg case control comparisons includes *T.b. gambiense* participants from Guinea and Côte d’Ivoire, Tbg case control comparison includes participants from *T.b. gambiense* endemic areas of Guinea, Côte d’Ivoire, Cameroon, DRC and North West Uganda. Tbr case control comparison includes participants from *T.b. rhodesiense* endemic areas of Malawi and South East and Central Uganda.

|  | Discovery Cohort |  |  | Validation Cohort |  |  |
| --- | --- | --- | --- | --- | --- | --- |
| Contrast | Cases | Control | Latent | Cases | Control | Latent |
| All Case Control | 958 | 1,089 | 0 | 570 | 1,033 | 0 |
| Tbg Case Control | 689 | 561 | 0 | 455 | 950 | 0 |
| Tbr Case Control | 269 | 528 | 0 | 115 | 83 | 0 |
| Tbg Case Latent | 417 | 0 | 94 | 393 | 0 | 69 |
| Tbg Control Latent | 0 | 245 | 94 | 0 | 280 | 69 |

Gemma was used to analyse each contrast listed in table 2 separately in a GWAS using the 1.64m genotyped SNP remaining after filtering. QQ plots indicated that the distributions of p values were as expected under a random model (Supplementary data Fig S1), Lambda values for all contrasts were near one (range 1.003 – 1.0071) indicating that there was little genomic inflation (Table S1).

SNP with a nominal P value < 10^−6^ in any of the five analyses were considered markers for candidate loci and were used to identify non-overlapping 200kb candidate regions (100kb either side of index SNP) in the five contrasts tested. Imputation against the 1000 genomes genotype panel was used to obtain 96k SNP within these candidate regions which were tested for association with phenotype in each of the five contrasts shown in Table 2. 88 SNP at 57 loci were genotyped in an additional 1,627 participants (Tables 1 and 2) to validate the candidate loci identified. After validation SNP rs76826738 in an intron of *SMOC2* had genome wide significant p value (2.95 x10^−8^) in the *T. b. gambiense* case control contrast (Tables 3&4, Figure 2). A further two loci around *NXN* and *NCKAP5* had suggestive associations (p < 10^−6^) after validation and one more was not validated but had a p < 10^−7^ in the primary analysis (Tables 3 & 4 and Fig 2).

**Table 3.**
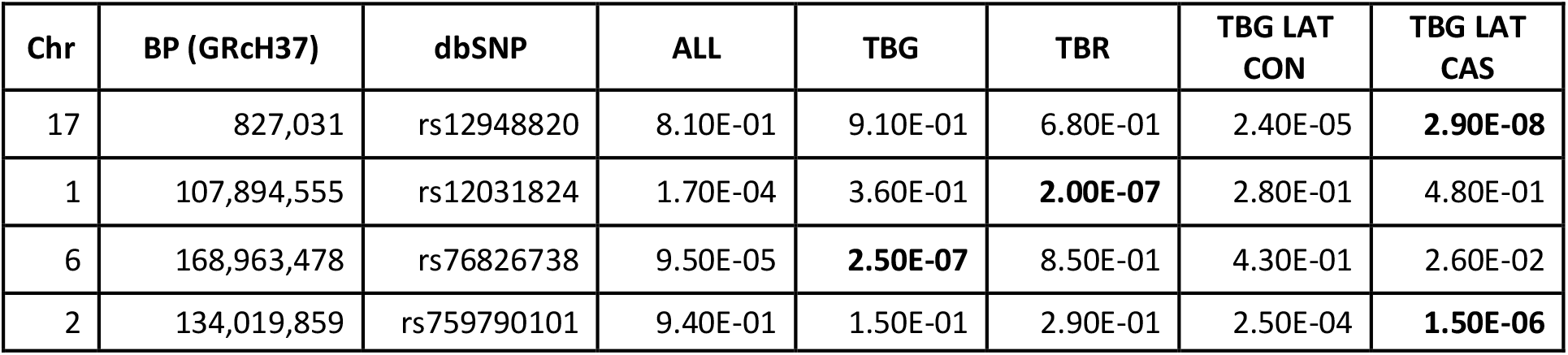
P values from the primary analysis for associations at loci that passed the genome wide threshold after the validation analysis. The genes listed are those in the 200kb regions centred on the peak SNP shown in the BP columns. The contrast with the smallest p value at each locus is highlighted in bold.

**Table 4.** P values, odds ratios, and minor allele frequency (MAF) for the three SNP which had p < 1 × 10^−6^ after meta-analysis. MAF observed in this study were very similar (±0.05) to those observed in the 1000 genomes data for Africa (Supplementary Data Table S2). Validation genotyping of the rs12031824 SNP failed.

| dbSnp | Genes | Contrast | MAF | OR | GWAS P | Validation P | Meta P |
| --- | --- | --- | --- | --- | --- | --- | --- |
| rs12948820 | TIMM22, NXN, ABR | TBG LAT CAS | 0.188 | 0.844 | 2.93E-08 | 0.039 | 2.32E-07 |
| rs12031824 | NTNG1 | TBR CAS CON | 0.178 | 0.851 | 2.04E-07 | n.d. | na |
| rs759790101 | NCKAP5 | TBG LAT CAS | 0.089 | 0.826 | 1.52E-06 | 0.009 | 3.41E-07 |
| rs76826738 | SMOC2 | TBG CAS CON | 0.115 | 1.176 | 2.50E-07 | 0.006 | 2.95E-08 |

**Figure 2.**
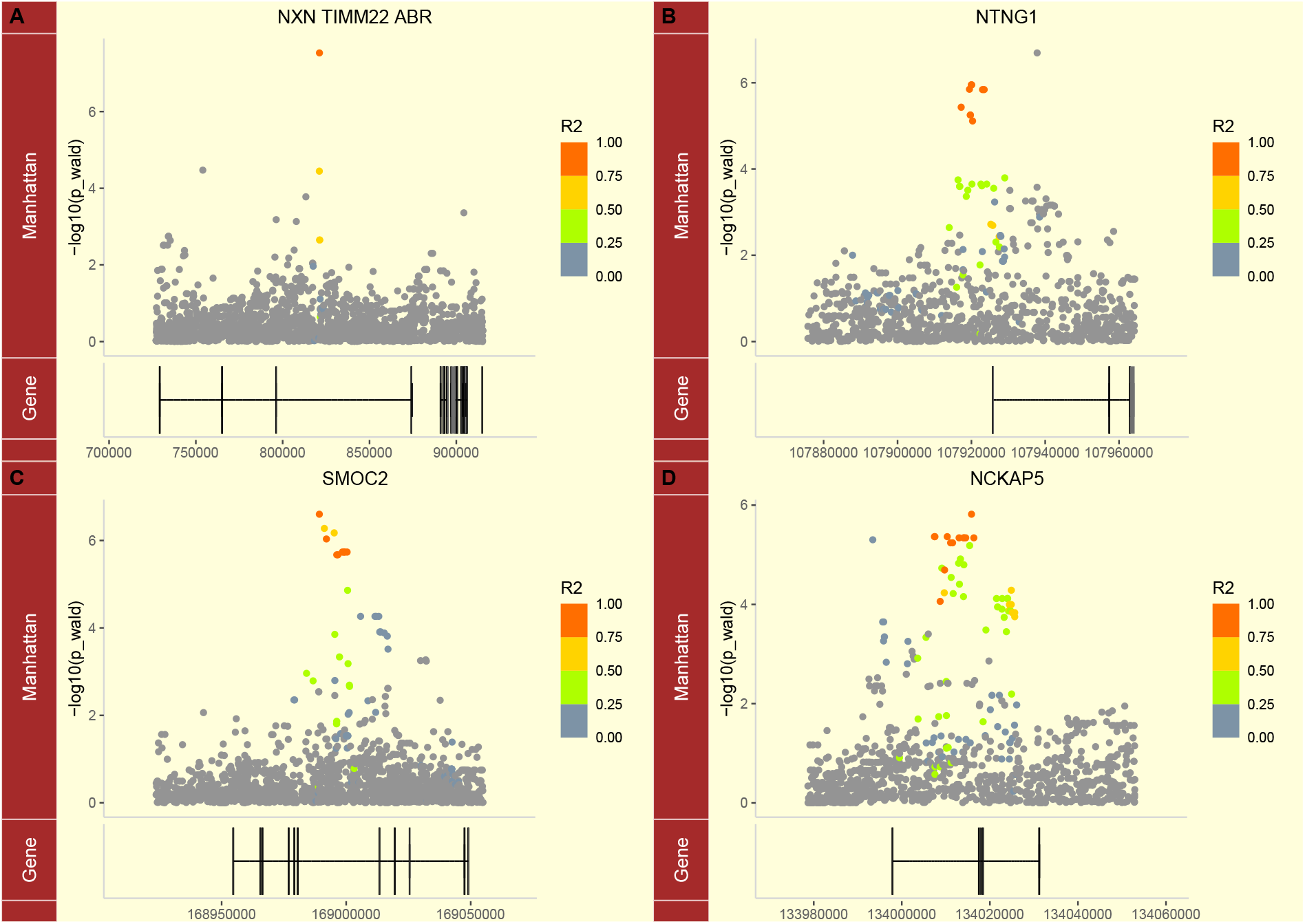
Region plots for top loci. Exons in genes in loci are shown below the Manhattan plot. Genes appear in the order listed in the title of the plot. Linkage to the index SNP is indicated by the colour.

The lead SNP at three of the four loci were members of large haplotypes that have consistent associations with infection (Fig 2). At the *NTNG1* and *NCKAP5* loci there appeared to be two distinct haplotypes associated with infection (Fig 2). The *NXN* QTL was the least well supported with only three linked SNP. The distribution of phenotypes between genotypes for the candidate SNP at these loci is shown in figure S2.

##### Associations of *APOL1* variants with HAT

*APOL1* has been intensively studied because two variants in this gene, designated G1 (rs73885319) and G2 (rs71785313) are only found in people with African ancestry and are associated with increased risk of chronic kidney disease (CKD). APOL1 confers protection against infection with *T. brucei brucei*. The two *T. brucei* subspecies that cause HAT have developed mechanisms to evade lysis by APOL1. The G1 and G2 variants that increase risk of CKD are assumed to have arisen in Africa to protect against HAT. The *APOL1* G2 variant is an indel and not represented on the H3Africa Consortium Array but genotypes were generated with a custom assay. Associations between HAT and *APOL1* variants are shown in Table 5. Notably the associations are population specific. G1 is only associated with latent infections in the West African population and not in case control contrasts. G2 is associated with latent infections in West Africa but also with a case control contrast but only in the *T.b. rhodesiense* infected population, in agreement with a previous candidate gene study (Cooper et al. 2017).

**Table 5.** Odds ratios and p values for association between *APOL1* genotypes associated with CKD in the five contrasts tested in the GWAS G1 is rs73885319 and G2 is rs71785313.

| Contrast | G1 OR | G1 p | G2 OR | G2 p |
| --- | --- | --- | --- | --- |
| All Case Control | 1.03 | 0.276 | 1.06 | 0.013 |
| Tbg Case Control | 1.05 | 0.094 | 1.00 | 0.936 |
| Tbr Case Control | 1.00 | 0.944 | 1.20 | 3.9E-06 |
| Tbg Case Latent | 0.88 | 4.1E-05 | 1.14 | 7.1E-05 |
| Tbg Control Latent | 0.89 | 0.003 | 1.18 | 7.0E-04 |

**Expression** of genes in the four candidate loci were determined in samples from DRC, Malawi and Guinea. The only genes that were associated with HAT and differentially expressed were *NXN* with a log2 fold change of 0.79 in skin of participants from Guinea (FDR = 0.0140) and TIMM22 (in the same QTL) with a log2 fold change of -0.38 in blood of participants from DRC (FDR = 0.0038).

## Discussion

The aim of our GWAS was to identify candidate loci that might regulate response to infection with *T. b. rhodesiense* or *T. b. gambiense*. These two parasite subspecies cause distinctive clinical syndromes with *T. b. rhodesiense* being associated with an acute disease, with progression to central nervous system involvement typically occurring within months, whilst *T. b. gambiense* infections can take years or decades to progress to CNS involvement. The TrypanoGEN initiative worked closely with the national control programmes of each country generating a valuable biobank (Ilboudo et al. 2017). We also sequenced the genomes of 298 participants from our study populations (Mulindwa et al. 2020) to contribute data to the H3Africa SNP chip initiative, which designed the chip that was used in the present study (Choudhury et al. 2020), to ensure that SNP that were rare outside Africa and therefore not included in commercial assays were included in our analysis.

The high diversity of African populations and the two distinct clinical syndromes associated with different parasite subspecies inevitably reduces the power of the GWAS. We addressed this problem by analysing the data by subpopulations and diseases. Although the corresponding reduction in numbers further reduces power, the increased homogeneity of the populations and diseases appears to have more than compensated for this since we found one genome wide significant associations with HAT and three further suggestive associations.

Two of the four loci with associations with HAT were for the cases vs latent contrast (Table 3) despite this contrast having the smallest number of samples. It is possible that this observation is an artefact of small sample size, however this is the contrast between the best-defined phenotypes. The cases and latents are defined by positive test reactions whereas all other contrasts include controls which are likely to include participants who are genetically susceptible but have not been exposed or participants who are genetically resistant and have either not been exposed or are susceptible but have self-cured. These participants with unknown status are likely reduce the power of tests for associations.

The region plots (Fig 2) show that the lead SNP at three of the QTL are members of large haplotypes which show a consistent pattern of association with disease and increase confidence that the associations are not due to technical error.

We also detected an association with APOL1 variants G1 and G2 which have been previously shown to regulate response to trypanosome infections (Kamoto et al. 2019; Cooper et al. 2017; Genovese et al. 2010) although the significance did not reach genome-wide levels. This gives us confidence that the observed associations we did detect at genome-wide significance levels are likely to have important impacts on the course of infection. The APOL1 variants had parasite subspecies specific associations with the G1 allele being associated with protection from progression from latent to active disease in *T.b. gambiense* infections and the G2 allele being associated with protection against *T.b. rhodesiense* HAT in Malawi and South East Uganda but not in Central Uganda (Cooper et al. 2017; Kamoto et al. 2019; Kimuda et al. 2018).

The only SNP to achieve genome wide significance after validation was rs76826738 in an intron in *SMOC2* for the comparison between *T.b. gambiense* cases and controls. It is not known if rs76826738 or another linked SNP is functional or how the functional variant modifies *SMOC2* expression or structure. SMOC2 is involved in the growth and differentiation of endothelial cells and may regulate the permeability of the endothelium (Maier, Paulsson, and Hartmann 2008; Schmidt et al. 2021).Trypanosomes need to transit the epithelia to leave and enter the bloodstream as well as to establish the initial infection and an increase in endothelial permeability might facilitate parasite invasion and dissemination.

*SMOC2* has also been associated with modifying the *APOL1* mediated risk of chronic kidney disease (CKD) (Chaudhary et al. 2022). Since *SMOC2* and *APOL1* appear to act together to increase the risk of kidney disease it is plausible that may also act together to reduce the risk of HAT.

NXN is a member of the thioredoxin family that catalyzes disulfide bond formation and isomerization and may be a master regulator of cellular redox homeostasis (Idelfonso-García et al. 2022). The lead SNP at the NXN locus had the least genetic supporting evidence since it appeared to be on a very small haplotype, however it had support from gene expression analysis in that increased expression of NXN in the skin was associated with protection against HAT.

## Conclusion

The TrypanoGEN GWAS has been remarkably successful in identifying four candidate QTL one of which was genome-wide significance after validation. These genes provide valuable entry points for further studies of the pathways regulating response to this deadly disease.

Despite the remarkable successes of the control programmes there is evidence of large reservoirs of asymptomatic infection which could lead to rapidly escalating outbreaks in adverse circumstances, as has happened before when wars have disrupted health care and farming in Uganda, Sudan, DRC and Angola. It is therefore essential to deepen our understanding of these diseases, particularly in the asymptomatic forms, represented in this study by the latent infections, in order to move from control to eradication. Our identification of loci associated with this latent phenotype are therefore of particular interest as they may lead to novel insights about this important reservoir population.

## Supporting information

Sopplementary Information

## Data Availability

All data produced in the present study are available upon reasonable request to the authors

## Acknowledgements

This work was supported by Human Heredity and Health in Africa (H3Africa) programme under Wellcome Trust grant number 099310/Z/12/Z and H3Africa grant number H3A-18-004. H3Africa is managed by the Science for Africa Foundation (SFA Foundation) in partnership with Wellcome, NIH and AfSHG. The views expressed herein are those of the author(s) and not necessarily those of the SFA Foundation and her partners.

We thank all the participants who donated samples to this project and the field work staff who assisted in the sample collection.

