## Supplementary material for "A Genome-Wide Association Study Reveals Novel Genetic Markers Associated with Human African Trypanosomiasis": Sopplementary Information

### Supplementary Information

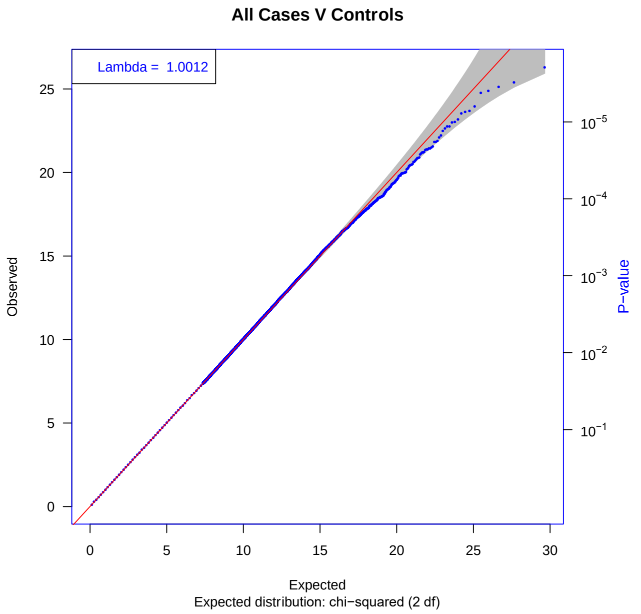


**Fig S1.** QQ plot of distribution of p values for all cases vs controls. The distribution was as expected under the null hypothesis of a uniform distribution of p values and the lambda of 1.0012 shows negligible evidence of genomic inflation.

| Contrast | Lambda |
|---|---|
| All Case Control | 1.0012 |
| Tbg Case Control | 1.0003 |
| Tbr Case Control | 1.0042 |
| WAF Case Latent | 1.0071 |
| WAF Control Latent | 1.0059 |

**Table S1.** Coefficients of genomic inflation (lambda) for the contrasts tested.

| Rs id | Location | TGEN_AF | AF | AFR_AF | AMR_AF | EAS_AF | EUR_AF | SAS_AF |
|---|---|---|---|---|---|---|---|---|
| rs12031824 | 1 107894555 | 0.162 | 0.1352 | 0.1339 | 0.2161 | 0.1448 | 0.0557 | 0.1513 |
| rs12948820 | 17 827031 | 0.225 | 0.3217 | 0.1997 | 0.2925 | 0.2857 | 0.4463 | 0.4162 |
| rs759790101 | 2:134019861 | | - | - | - | - | - | - |
| rs76826738 | 6:168963478 | 0.108 | 0.0326 | 0.1157 | 0.0144 | 0 | 0 | 0 |

**Table S2** Minor allele frequencies observed in the current study (TGEN_AF) compared with allele frequencies observed in 1000 genomes data. Frequencies were very similar in the two data sets. AF global MAF, AFR_AF African MAF; EAS_AF East Asian MAF; EUR_AF European AF; SAS_AF South Asian MAF. Note rs759790101 is not represented in 1000 genomes data and the MAF at the SMOC2 locus on chr 6 rs76826738 is < 2% in all populations except Africans.

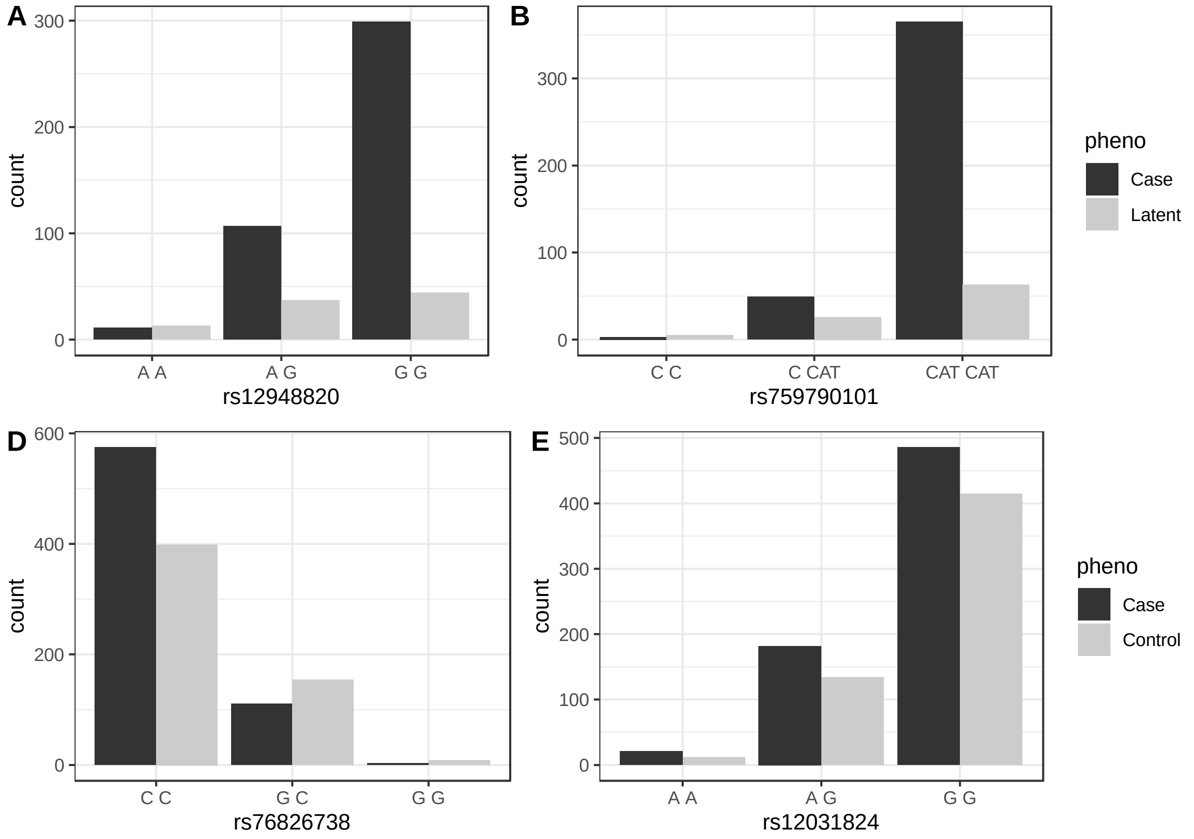


**Figure S2**. The number of participants with each phenotype for each genotype of each candidate SNP. The counts were obtained for the contrasts shown in Table 4.
